# Elevated mortality among people experiencing homelessness with COVID-19

**DOI:** 10.1101/2021.03.05.21253019

**Authors:** Kathryn M. Leifheit, Lelia H. Chaisson, Jesus Alejandro Medina, Rafik Wahbi, Chelsea L. Shover

**Affiliations:** UCLA Fielding School of Public Health, Department of Health Policy & Management; University of Illinois at Chicago, Department of Medicine, Division of Infectious Diseases; UCLA David Geffen School of Medicine; UCLA Fielding School of Public Health, Department of Community Health Sciences; UCLA David Geffen School of Medicine, Department of Medicine, Division of General Internal Medicine and Health Services Research

## Abstract

We reviewed publicly available data from major U.S. health jurisdictions to compare case fatality rates in people experiencing homelessness (PEH) to the general population. Case fatality among PEH was 1.3 times (95% CI 1.1, 1.5) that of the general population, suggesting that PEH should be prioritized for vaccination.

## Introduction

People experiencing homelessness (PEH) are highly vulnerable to COVID-19 due to numerous factors including increased risk of SARS-CoV-2 exposure and infection. Lacking permanent shelter, PEH are less able to mitigate exposures through social distancing, often living doubled up in the homes of friends and family, in crowded shelter facilities, or in densely populated encampments. Recommended strategies to prevent infection may be challenging for PEH: access to high-quality masks may be limited and inconsistent access to running water makes hand hygiene challenging. Through the course of the pandemic, health authorities have reported high seroprevalence among PEH^1^ and large outbreaks at homeless shelters and encampments.^2–5^ Relatedly, research has linked evictions to community-level COVID-19 risk.^6,7^

Once infected, PEH may be at elevated risk of COVID-19 morbidity and mortality. Homelessness takes a dramatic toll on health. Researchers have estimated life expectancies among the chronically homeless to be as low as 42-52 years.^8^ Weathering homelessness means that PEH develop age-related health problems decades earlier than their housed counterparts.^9,10^ Many of these conditions may increase risk for severe illness and death from COVID-19. For example, the prevalence of chronic obstructive pulmonary disease among PEH may be two to three times that of the general population.^10–12^ Based on this underlying risk, a modeling study projected that 4% of the U.S. homeless population would require hospitalization due to COVID-19 infections and that nearly 1% would die.^13^

Despite substantial vulnerability to COVID-19 among PEH, few data exist to gauge the impact of homelessness on COVID-19 outcomes. We therefore reviewed publicly available data in U.S. health jurisdictions to compare case fatality rates among PEH to population-wide case fatality rates.

## Methods

We systematically searched for PEH-specific COVID-19 case and mortality data on websites from public health agencies with jurisdiction over the 25 most populous U.S. counties (n=25), the 25 most populous U.S. cities (n=12 additional), and the most populous city in each state (n=39 additional). Data were collected from February 19^th^ – 28^th^, 2021.

If PEH-specific counts were available, we extracted the most recent cumulative case and death counts for PEH, as well as jurisdiction-wide case and death counts. Where possible, we also extracted data by demographic factors such as age.

For each jurisdiction, we calculated PEH case fatality rates (CFRs, defined as deaths per 100 cases) and jurisdiction-wide CFRs, and compared these via relative risk (a ratio of PEH CFR over jurisdiction-wide CFR). We then summarized CFRs and relative risk estimates across all jurisdictions as a population-weighted average. If a jurisdiction published age-stratified PEH case and death counts, we calculated age-specific CFRs and relative risk.

## Results

Of the 76 jurisdictions reviewed, 7 (9.2%) with data on PEH were included. In these jurisdictions, the CFR for PEH ranged from 0.3% to 4.8%, compared to jurisdiction-wide CFRs that ranged from 0.6% to 2.5% (Figure 1a, relative risks ranging from 0.3 to 2.2). Across jurisdictions, the overall CFR was 2.1% (95% CI 1.8, 2.3) among PEH and 1.6% (CI 1.6, 1.6) for the general population (relative risk 1.3 (CI 1.1, 1.5)).

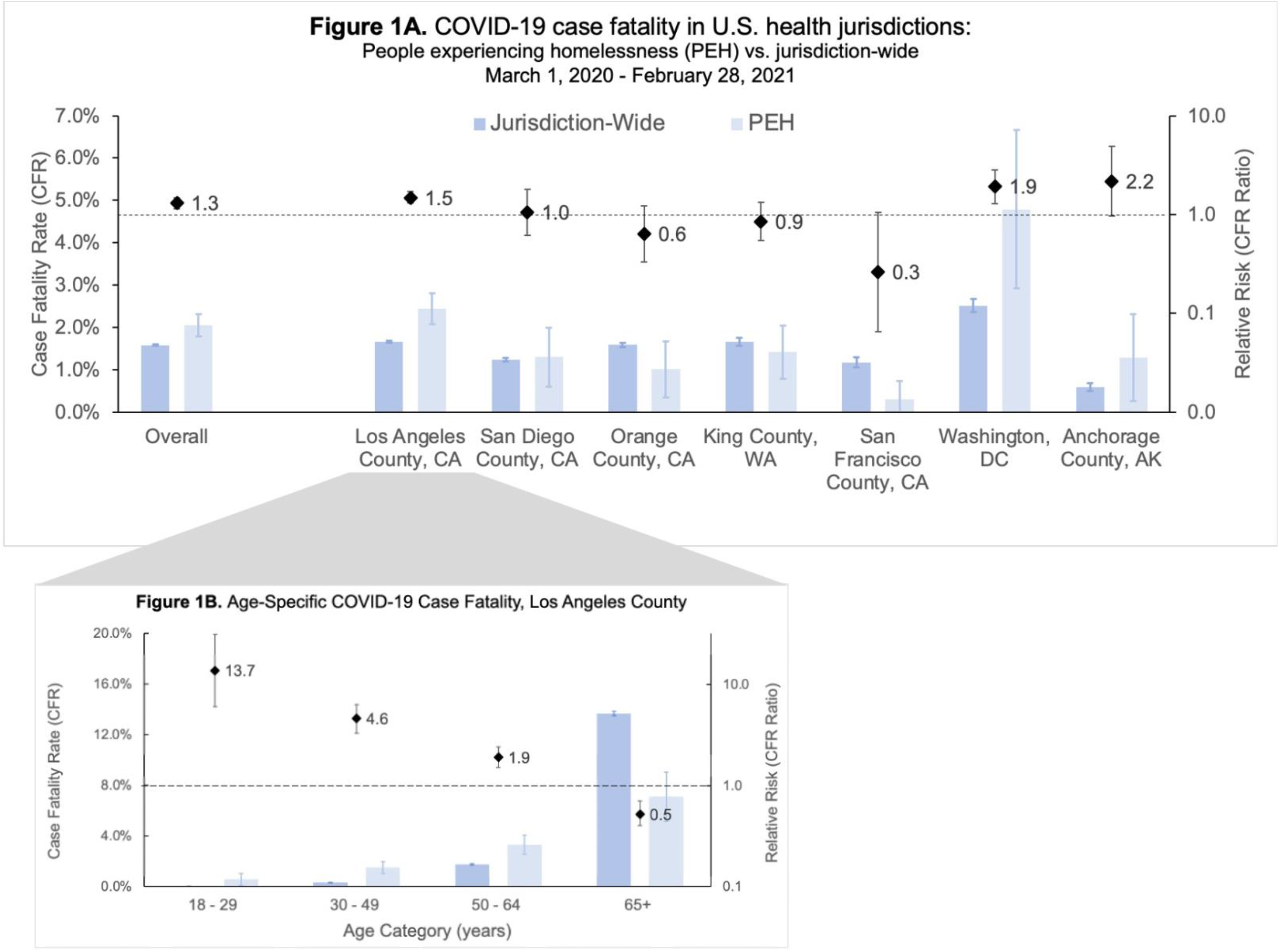

Only Los Angeles County stratified PEH data by additional demographic characteristics. We compared age-specific CFRs between PEH and the county population (Figure 1b), and found CFRs were significantly increased for PEH under 65 compared to the county population, with relative risks of 13.7 (95% CI 6.0, 31.1), 4.6 (3.3, 6.3), and 1.9 (1.5, 2.4) for 18-29, 30-49, and 50-64 year-olds, respectively. Among adults aged 65 and over, this trend was reversed, with a relative risk of 0.5 (0.4, 0.7).

## Discussion

Our review identified few jurisdictions with publicly available information on COVID-19 cases and mortality among PEH; however, the available data reveal significantly elevated overall and age-specific COVID-19 mortality among PEH compared to the overall population. Individuals identified as experiencing homelessness at the time of COVID-19 diagnosis had a 30% higher risk of death than the overall population, underscoring the need to prioritize PEH for COVID-19 prevention interventions, including vaccination. Because this increased risk was especially pronounced in younger age groups, age-based vaccine eligibility criteria may be inappropriate for this medically vulnerable population.

Expanding vaccination eligibility to all PEH, regardless of age or presence of comorbidities, would prevent COVID-19 mortality, while also streamlining vaccine distribution. The CDC prioritized residents of long-term care facilities in Phase 1A of vaccine distribution because congregate living and medical vulnerability placed residents at increased risk of COVID-19 infection and death, respectively.^14^ The same reasoning applies to PEH. Location-based eligibility would enable jurisdictions to more efficiently leverage homeless outreach and healthcare personnel. For example, vaccinating all consenting adults at one shelter or encampment and then moving to the next reduces need for repeat visits to deliver vaccine to only a small portion of eligible individuals at a time.

We found substantial heterogeneity in case fatality and CFR ratios across jurisdictions, likely reflecting local differences in COVID-19 dynamics, surveillance and testing practices, homeless services, and reporting. Careful evaluation of local COVID-19 prevention and control programs is important to identify effective strategies to reduce the burden of COVID-19 in PEH. Interestingly, we found that PEH cases over 65 years had a lower risk of COVID-19 mortality than cases in Los Angeles’ overall over-65 population. The most likely explanation for this finding is survivor bias: on average, PEH who survive to age 65 without shelter may be less frail than older adults in the general population. Given low life expectancy among PEH,^8^ we also hypothesize differing age distributions within this stratum, with fewer PEH aged 80 and over. Importantly, however, older PEH remained at substantially elevated mortality risk compared with younger PEH.

There are several limitations to our study. Because so few jurisdictions published PEH-specific data, the generalizability of our estimates may be limited. Since available PEH data were rarely disaggregated, we were not able to account for underlying differences such as age structure in our comparisons of PEH versus jurisdiction-wide cases. Additionally, PEH status is prone to misclassification and differential classification across jurisdictions. Specifically, we expect that PEH not accessing homeless services would be misclassified as not PEH. Furthermore, COVID-19 screening programs by homeless services and health departments may increase detection of mild COVID-19 cases and asymptomatic SARS-CoV-2 infections among PEH compared with the general population. Importantly, exposure misclassification and differential surveillance would likely lead to underestimation of the degree to which homelessness increases COVID-19 case fatality, strengthening our conclusion that PEH are a highly vulnerable population in need of effective interventions to prevent COVID-19.

The paucity of COVID-19 data specific to PEH represents a limitation not just for our study, but for public health surveillance as a whole. Disaggregated data allow for development of targeted public health interventions that prioritize high-risk populations for case finding and vaccination. Moreover, these data allow for monitoring of health inequities, documenting the degree to which PEH are disproportionately burdened by COVID-19. Moving forward, it is essential that data on homelessness be made available to understand the degree to which structural failures such as insufficient housing have compromised U.S. pandemic preparedness. Routinely collecting housing status as part of disease surveillance may pose logistical challenges in some jurisdictions, as clinics, testing sites, laboratories, and coroners would need to coordinate carefully. Nonetheless, we see this as an important step for the control of COVID-19 and other infectious diseases in this high risk and underserved population.

As COVID-19 vaccines bring new signs of hope, we urge public agencies to prioritize PEH for vaccination. Past outbreaks and seroprevalence studies have shown that exposure and infection risk are extremely high among PEH. Our analyses suggest that COVID-19 may also be particularly deadly for this population, independent of age. Vaccinating PEH has the potential to prevent deaths, promote health equity, and reduce community transmission of COVID-19 in many of the country’s hardest hit cities.^15^

## Data Availability

Data were extracted from public-facing health department websites. Our data extraction form is available upon request.

## References

1. Roederer T, Mollo B, Vincent C, et al. Seroprevalence and risk factors of exposure to COVID-19 in homeless people in Paris, France: a cross-sectional study. Lancet Public Heal. 2021;2667(21):1–3. doi:10.1016/s2468-2667(21)00001-3

2. Maxmen A. Coronavirus is spreading under the radar in US homeless shelters. Nature. 2020;581(7807):129–130. doi:10.1038/d41586-020-01389-3

3. Baggett TP, Keyes H, Sporn N, Gaeta JM. Prevalence of SARS-CoV-2 Infection in Residents of a Large Homeless Shelter in Boston. JAMA - J Am Med Assoc. Published online 2020. doi:10.1001/jama.2020.6887

4. Imbert E, Kinley PM, Scarborough A, et al. Coronavirus Disease 2019 Outbreak in a San Francisco Homeless Shelter. Clin Infect Dis. Published online 2020. doi:10.1093/cid/ciaa1071

5. Kendall M. Coronavirus outbreak: 11 test positive at Oakland homeless camp. The Mercury News.

6. Sheen J, Nande A, Walters EL, et al. The effect of eviction moratoriums on the transmission of SARS-CoV-2. medRxiv. Published online January 1, 2020:2020.10.27.20220897. doi:10.1101/2020.10.27.20220897

7. Leifheit K, Linton S, Raifman J, et al. Expiring eviction moratoriums and COVID-19 incidence and mortality. SSRN. Published 2020. https://papers.ssrn.com/sol3/papers.cfm?abstract_id=3739576

8. O’Connell J. Premature Mortality in Homeless Populations: A review of the literature. N Engl J Med. Published online 2005.

9. Adams J, Rosenheck R, Gee L, Seibyl CL, Kushel M. Hospitalized younger: A comparison of a national sample of homeless and housed inpatient veterans. J Health Care Poor Underserved. Published online 2007. doi:10.1353/hpu.2007.0000

10. Brown RT, Kiely DK, Bharel M, Mitchell SL. Geriatric syndromes in older homeless adults. J Gen Intern Med. Published online 2012. doi:10.1007/s11606-011-1848-9

11. Snyder LD, Eisner MD. Obstructive lung disease among the urban homeless. Chest. Published online 2004. doi:10.1378/chest.125.5.1719

12. Landis SH, Muellerova H, Mannino DM, et al. Continuing to confront COPD international patient survey: Methods, COPD prevalence, and disease burden in 2012-2013. Int J COPD. Published online 2014. doi:10.2147/COPD.S61854

13. Culhane D, Treglia D, Steif K, Kuhn R, Byrn T. Estimated Emergency and Observational/Quarantine Capacity Need for the US Homeless Population Related to COVID-19 Exposure by County; Projected Hospitalizations, Intensive Care Units and Mortality.; 2020.

14. Dooling K, McClung N, Chamberland M, et al. The Advisory Committee on Immunization Practices’ Interim Recommendation for Allocating Initial Supplies of COVID-19 Vaccine — United States, 2020. MMWR Morb Mortal Wkly Rep. Published online 2020. doi:10.15585/mmwr.mm6949e1

15. Bibbins-Domingo K, Petersen M, Havlir D. Taking Vaccine to Where the Virus Is—Equity and Effectiveness in Coronavirus Vaccinations. JAMA Heal Forum. 2021;2(2):e210213–e210213. doi:10.1001/jamahealthforum.2021.0213

